# Early outcomes of tocilizumab in adults hospitalized with severe COVID-19 - The Vall d’Hebron COVID-19 prospective cohort study

**DOI:** 10.1101/2020.05.07.20094599

**Authors:** Adrián Sánchez-Montalvá, Júlia Sellarés-Nadal, Juan Espinosa-Pereiro, Nuria Fernández-Hidalgo, Santiago Pérez-Hoyos, Fernando Salvador, Xavier Durà, Marta Miarons, Andrés Antón, Simeón Eremiev-Eremiev, Abiu Sempere-González, Arnau Monforte-Pallarés, Pau Bosch-Nicolau, Salvador Augustin, Júlia Sampol, Alfredo Guillén-del-Castillo, Benito Almirante

**Affiliations:** Infectious Diseases Department, Vall d’Hebron University Hospital, Universitat Autònoma de Barcelona, Barcelona, Spain; Statistic Unit, Vall d’Hebron Institute of Research, Barcelona, Spain; Pharmacy Department, Vall d’Hebron University Hospital, Universitat Autònoma de Barcelona, Barcelona, Spain; Microbiology Department, Vall d’Hebron University Hospital, Universitat Autònoma de Barcelona, Barcelona, Spain; Hepatology Department, Vall d’Hebron University Hospital, Universitat Autònoma de Barcelona, Barcelona, Spain; Pneumology Department, Vall d’Hebron University Hospital, Universitat Autònoma de Barcelona, Barcelona, Spain; Internal Medicine Department, Vall d’Hebron University Hospital, Universitat Autònoma de Barcelona, Barcelona, Spain; International Health Program of the Catalan Institut of Health (PROSICS), Barcelona, Spain; Tropical Medicine Spanish Research Network (RICET), Madrid, Spain; Infectous Diseases Spanish Research Network (REIPI), Madrid, Spain

## Abstract

**Background:** Modulation of the immune system to prevent lung injury is being widely used against the new coronavirus disease (COVID-19) despite the scarcity of evidence.

**Methods:** We report the preliminary results from the Vall d’Hebron prospective cohort study at Vall d’Hebron University Hospital, in Barcelona (Spain), including all consecutive patients who had a confirmed infection with the severe acute respiratory syndrome coronavirus-2 (SARS-CoV-2) and who were treated with tocilizumab until March 25^th^. The primary endpoint was mortality at 7 days after tocilizumab administration. Secondary endpoints were admission to the intensive care unit, development of ARDS and respiratory insufficiency among others.

**Results:** 82 patients with COVID-19 received at least one dose of tocilizumab. The mean (± SD) age was 59.1 (19.8) years, 63% were male, 22% were of non-Spanish ancestry, and the median (IQR) age-adjusted Charlson index at baseline was 3 (1-4) points. Respiratory failure and ARDS developed in 62 (75.6%) and 45 (54.9%) patients, respectively. Median time from symptom onset to ARDS development was 8 (5-11) days. The median time from symptom onset to the first dose of tocilizumab was 9 (7-11) days. Mortality at 7 days was 26.8%. Hazard ratio for mortality was 3.3; 95% CI, 1.3 to 8.5 (age-adjusted hazard ratio for mortality 2.1; 95% CI, 0.8 to 5.8) if tocilizumab was administered after the onset of ARDS.

**Conclusion:** Time from lung injury onset to tocilizumab administration may be critical to patient recovery. Our preliminary data could inform bedside decisions until more data from clinical trials becomes available.

**Summary of the article’s main point:** In patient with COVID-19 and lung injury, time from lung injury onset to tocilizumab administration may be critical to patient recovery. Early administration of host-directed therapies may improve patient outcome.

## INTRODUCTION

Coronavirus disease 2019 (COVID-19) is a novel illness caused by severe acute respiratory syndrome coronavirus-2 (SARS-CoV-2). It was first reported in December 2019 in Hubei province, China.^1^ Since then, SARS-CoV-2 has rapidly spread worldwide. According to the World Health Organization (WHO), as of July 8^th^, 2020, there have been 11.669.259 laboratory-confirmed cases and 539.906 deaths.^2^ The crude case fatality rate, estimated to be between 2.3% and 3.3%, is highly dependent on age and underlying conditions.^3,4^ Death is mainly due to respiratory failure caused by an acute respiratory distress syndrome (ARDS). As the pathophisiology behind lung injury is progressively elucidated, several therapies have been proposed on the basis of pre-clinical studies and the previous experience with the severe acute respiratory syndrome coronavirus (SARS-CoV) and the Middle East Respiratory Syndrome Coronavirus (MERS-CoV).^5,6^ Many of them are being used off-label in a desperate attempt to improve patient outcomes, including antiviral therapies, coagulation-modifying drugs and immune-modulating therapies.^7–11^In COVID-19, an excessive immune response inducing disproportionate release of cytokines and hyperinflammation has been proposed as a cause for the lung damage, mimicking a secondary haemophagocytic lymphohistiocytosis.^12^ Host-directed therapies have immune-modulating properties with higher precision than steroids and other immune-modulating therapies.^13^ Tocilizumab is a humanized monoclonal antibody that inhibits interleukin-6 (IL-6) receptor with a well-known safety profile and is approved for the treatment of rheumatoid arthritis and, since 2017, the treatment of chimeric antigen receptor (CAR) T cell-induced severe or life-threatening cytokine release syndrome (CRS).^14,15^ It has been used with promising results in small retrospective cohort studies of SARS-CoV-2 infected patients in China.^9,16^ The efficacy of other host-directed therapies targeting hyperinflammation is being assessed under randomized clinical trial conditions for severe SARS-CoV-2 infected patients.^17,18^

However, a proper characterization of the subset of patients who will benefit most from host-directed therapy and defining the precise timing for host-directed therapies administration has not yet been performed and is critical to allocate limited drug stocks and reduce COVID-19 associated mortality. We aim to describe a prospective cohort of SARS-CoV-2 infected patients treated with tocilizumab and define risk factors associated with poor outcome.

## METHODS

### Study setting and population

The Vall d’Hebron COVID-19 Prospective Cohort Study includes all consecutive adult patients (≥ 18 years old) treated for COVID-19 at Vall d’Hebron University Hospital, a 1100-bed public tertiary care hospital in Barcelona, Spain. For this study we selected the subgroup of patients with laboratory-confirmed COVID-19 and radiologically confirmed pneumonia who received at least one dose of tocilizumab. Identification and inclusion of patients receiving tocilizumab was performed from the Pharmacy Department registry.

### Standard of care and tocilizumab administration criteria

At admission, all patients were initially evaluated with chest radiography and blood tests including complete cell count, coagulation studies, biochemistry and inflammatory parameters. Treatment with lopinavir/ritonavir, azithromycin and hydroxychloroquine was initiated according to Vall d’Hebron University Hospital protocol. Tocilizumab was considered as additional treatment in patients with the following criteria: 1) respiratory failure defined as a ratio of arterial oxygen tension to fraction of inspired oxygen (PaO2/FiO2 ratio) of <300, a ratio of arterial oxygen saturation measured by pulse oximetry to fraction of inspired oxygen (SpO2/FiO2 ratio) of <315 or pO2 <60mmHg or oxygen saturation measured by pulse oximetry less than 90% when breathing room air or rapidly progressive clinical worsening according to treating physician and 2) interleukin-6 (IL-6) levels >40pg/mL (reference 0-4.3pg/mL) or a D-dimer levels > 1500 ng/mL (reference 0-243 pg/mL). Two dosing regimens based on weight were considered for tocilizumab. Patients over 75kg received 600mg, otherwise 400mg was the preferred dose. A second dose was considered in patients with a poor early response. Patients with liver enzymes (aspartate aminotransferase and alanine aminotransferase) 5 times over the upper limit of normality or concomitant severe bacterial infection were not eligible for tocilizumab treatment.

### Data sources

Data were collected retrospectively from the medical charts of tpatients from the 13^th^ of March, 2020 to the 18^th^ of March, 2020, when the protocol was submitted to the institutional review board, and prospectively thereafter. Inclusion and follow-up are still ongoing. The cut-off data for inclusion in this sub-study was the 25^th^ of March, 2020. All patients included were followed for at least 7 days. The institutional review board provided ethical clearance (local review board code number: PR(AG)183/2020). Patients were asked for an oral consent. The institutional review board granted an informed consent waiver if patients were unable to give oral consent. Written consent was waived because of the crisis context and concerns about safety when introducing a physical support for the consent in the isolation areas.

A Laboratory-confirmed case was defined as a patient with a real-time reverse-transcriptase-polymerase-chain-reaction (RT-PCR) SARS-CoV-2 positive result in any respiratory sample (nasopharyngeal swab, sputum, bronchoalveolar lavage or aspirate, tracheal aspirate).

We collected sociodemographic characteristics, past medical history, Charlson comorbidity score, concomitant medication, current therapy, adverse drug events, blood test results, imaging studies, microbiological tests other than SARS-CoV-2 RT-PCR on respiratory samples when available, and supportive measures needed. Vital signs, symptoms and physical examination were evaluated on admission, at 48h and weekly during hospital admission. Laboratory, microbiology and imaging studies were performed on admission and thereafter according to the clinical care needs of each patient. Laboratory assessments consisted of a complete blood count, coagulation testing including D-dimer measurement, liver and renal tests, electrolyte profile, and inflammatory profile including C-reactive protein, fibrinogen, ferritin and IL-6. All radiographs were reviewed by the investigators and computed tomography (CT) scans were recorded according to the radiology department reports. The COVID-19 severity was measured with the CURB-65 scale for community acquired pneumonia and other scales.^19,20^ Data was recorded in the Research Electronic Data Capture software (REDCap, Vanderbilt University).

### Laboratory confirmation

From the onset of the outbreak until 15^th^ of March the microbiological diagnosis was based on a homebrew RT-PCR assay targeting two viral targets (N1 and N2) in the viral nucleocapsid (N) gene and one in the envelope (E) gene of SARS-CoV-2, as well as the human RNase P (RP) gene as an internal control of the whole process, according to the CDC and ECDC Real-Time RT-PCR Diagnostic Panels with minor modifications.^21^ Since March 15^th^, commercial Allplex™ 2019-nCoV multiplex RT-PCR (Seegene, South Korea) were used for the detection of three viral targets (E; N; and, RNA-dependent RNA polymerase, RdRp) and an internal control. First SARS-CoV-2 laboratory-confirmations were confirmed by RdRp sequencing.^22,23^ Total nucleic acids (DNA/RNA) were extracted from respiratory specimens using NucliSENS easyMAG (BioMerieux, France) and STARMag Universal Cartridge Kit (Seegene, South Korea) according to the manufacturer’s instructions. All microbiological procedures were carried out in the laboratory under Biosafety Level 2 conditions.

### Study outcomes

The primary simple endpoint was defined as death at 7 days after first dose of tocilizumab. Secondary outcomes were admission to Intensive Care Unit (ICU), acute Respiratory Distress Syndrome (ARDS) and respiratory insufficiency. We also assessed acute myocardial infarction, septic shock, acute kidney injury and secondary infections. Berlin criteria for the ARDS were adapted, as many of the patients did not have an available arterial O2 pressure due to the overwhelming volume of admitted patients that precluded us from performing arterial blood samples on all patients. Instead, we used oxygen saturation by pulse oximetry and its correlation to the inspired fraction of oxygen (SpO2/FiO2 ratio < 315).^24,25^ One patient died a few hours after receiving tocilizumab and was excluded from the primary endpoint analysis.

### Statistical analysis

Continuous variables were expressed as mean and standard deviation or medians and interquartile range, as appropriate. Categorical variables were summarized as absolute number and percentages. Comparisons among groups was performed with Chi squared test and Fisher’s test for categorical variables; and Student’s T test and Mann-Whitney U test for continuous variables. Box plots and bar plots are also provided for some associations. Mortality in the cohort was described with the use of Kaplan-Meier analysis. Tests were considered significant when the two-tailed p-value was <0.05. We did not correct for multiple comparisons; hence, the widths of the confidence intervals should not be interpreted as definitive for the associations with the outcomes. Association between time to tocilizumab administration and mortality were assessed by means of Cox proportional hazards regression. Missing urea and bilirubin levels on admission were assumed normal for CURB-65 and SOFA calculation; no other imputation was made for missing data. Analysis was performed with Stata 15.1 software (StataCorp).

### Study oversight

The study was designed and conducted by the investigators from the Vall d’Hebron COVID-19 Prospective Cohort Study. No specific funding was provided to conduct the study. Data were collected, debugged, analysed and interpreted by the authors. All the authors reviewed the manuscript and vouch for the accuracy and completeness of the data and for the adherence of the study to the protocol.

## RESULTS

### Demographic and clinical characteristics

Since the onset of the COVID-19 outbreak until March 25^th^, 3242 respiratory-derived samples have been requested from our institution for COVID-19 diagnosis. Samples were requested from the emergency room and hospital wards, as well as from the health care worker surveillance strategy plan. From them, 941 were positive (29%). During this period, 82 SARS-CoV-2 infected patients received at least one dose of tocilizumab. The mean (±SD) age was 59.1 (±19.8) years (95% Confidence Interval 54.8-63.5). Fifty-two patients were male (63.5%). Eighteen (21.9%) patients were born abroad, 13 (16.1%) in Latin America, 3 (3.7%) in Eastern Europe and 2 (2.4%) in North Africa. The mean (±SD) duration of symptoms before hospital admission was 6.7 (±4.4) days. Fever and cough were the main symptoms on admission, occurring in 75 (91.5%) and 71 (86.6%) cases respectively.

Thirty-three (40.3%) patients were former or active tobacco smokers. Coexisting conditions were as follows: 32 (39.0%) had hypertension, 19 (23.5%) had lung diseases (2 (2.4%) asthma, 6 (7.3%) chronic obstructive pulmonary disease among others), 17 (20.7%) had obesity, 16 (19.5%) had diabetes mellitus, 11 (13.6%) had chronic kidney disease, 5 (6.1%) a history of cardiac failure, 1 (1.2%) had cirrhosis. Ten (12.5%) patients were immunosuppressed because of different conditions. Seventy-seven (95.1%) patients had a Barthel scale index of 100 points previous to hospital admission. Table 1 shows demographic and clinical characteristics at baseline.

**Table 1.**
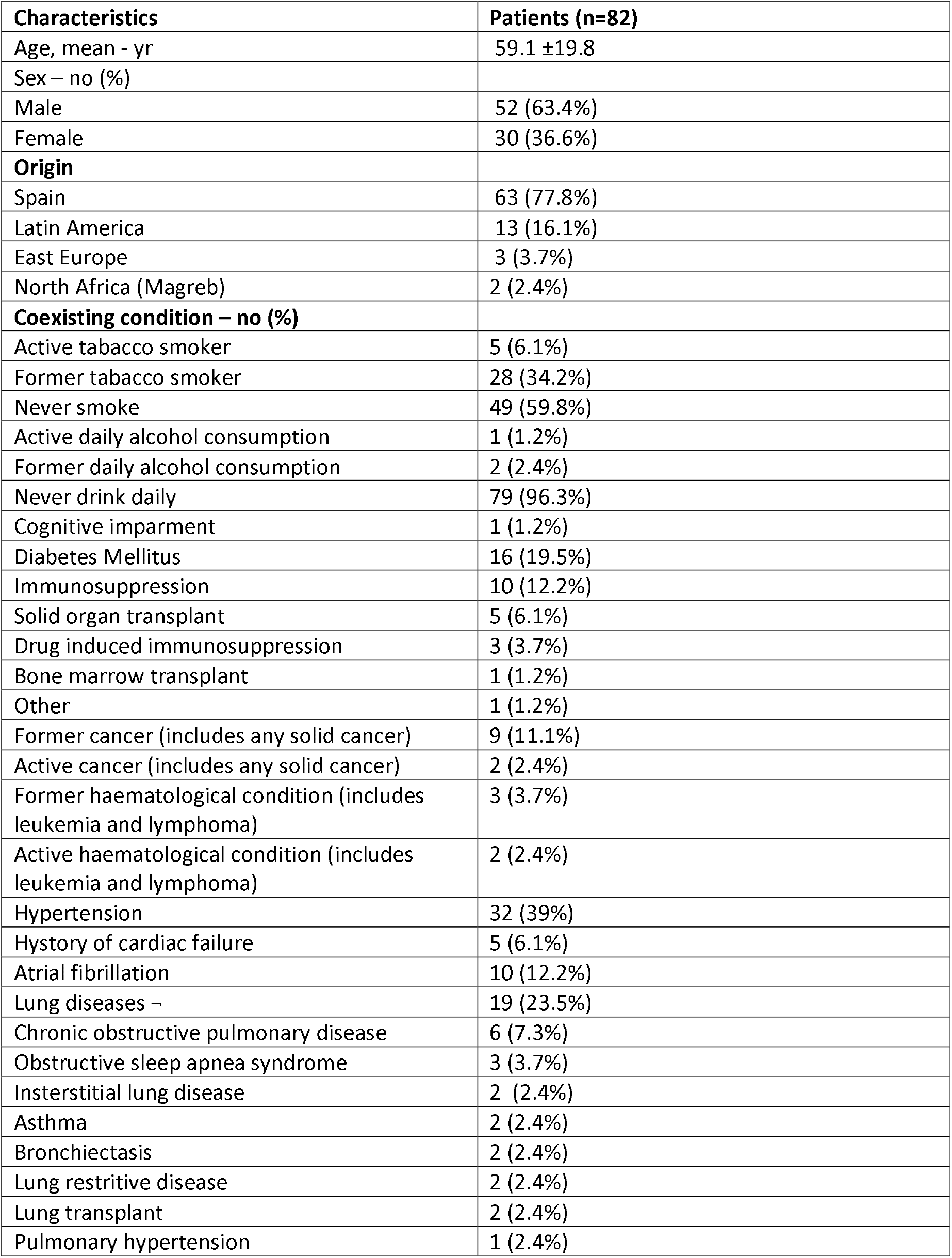
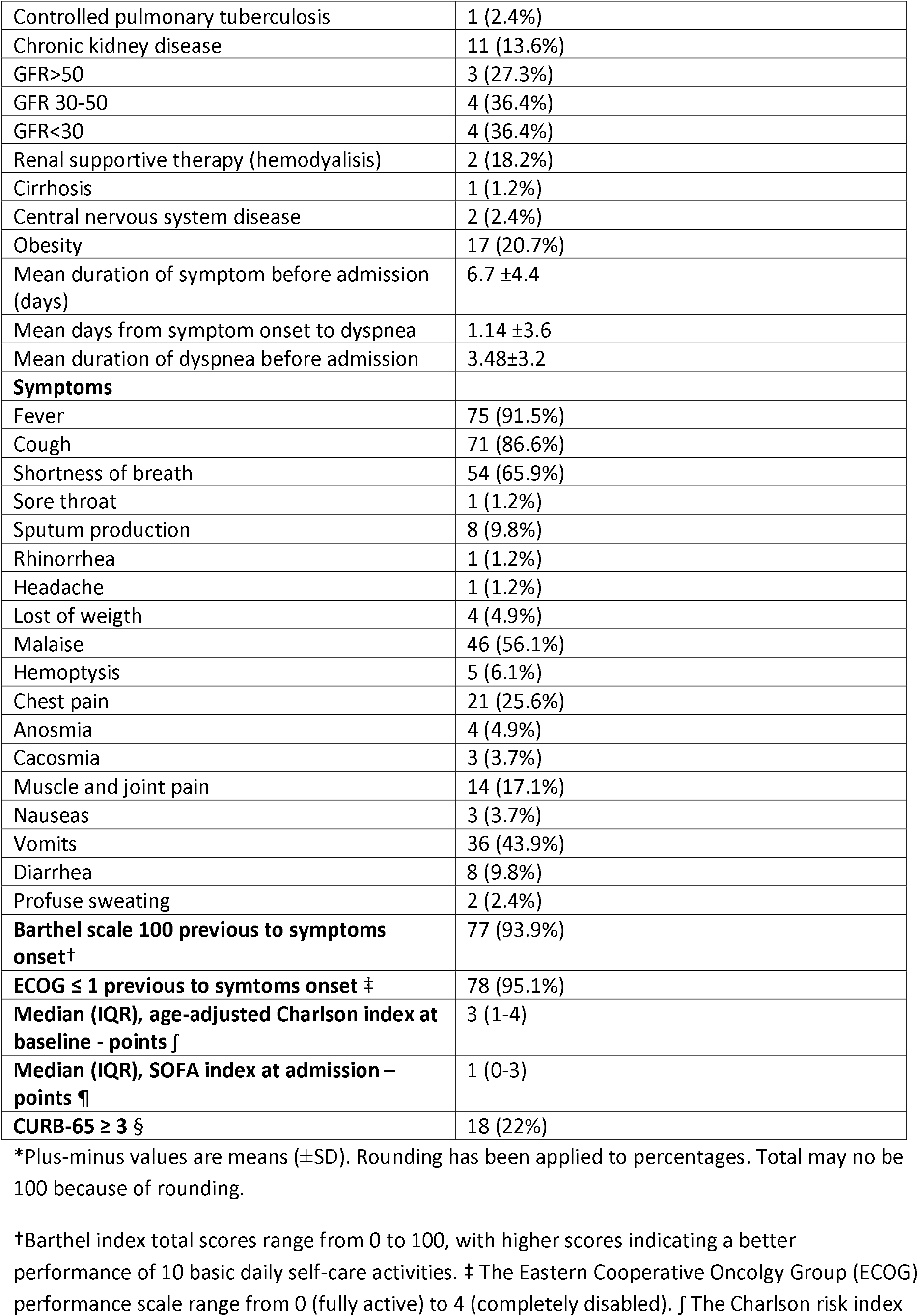
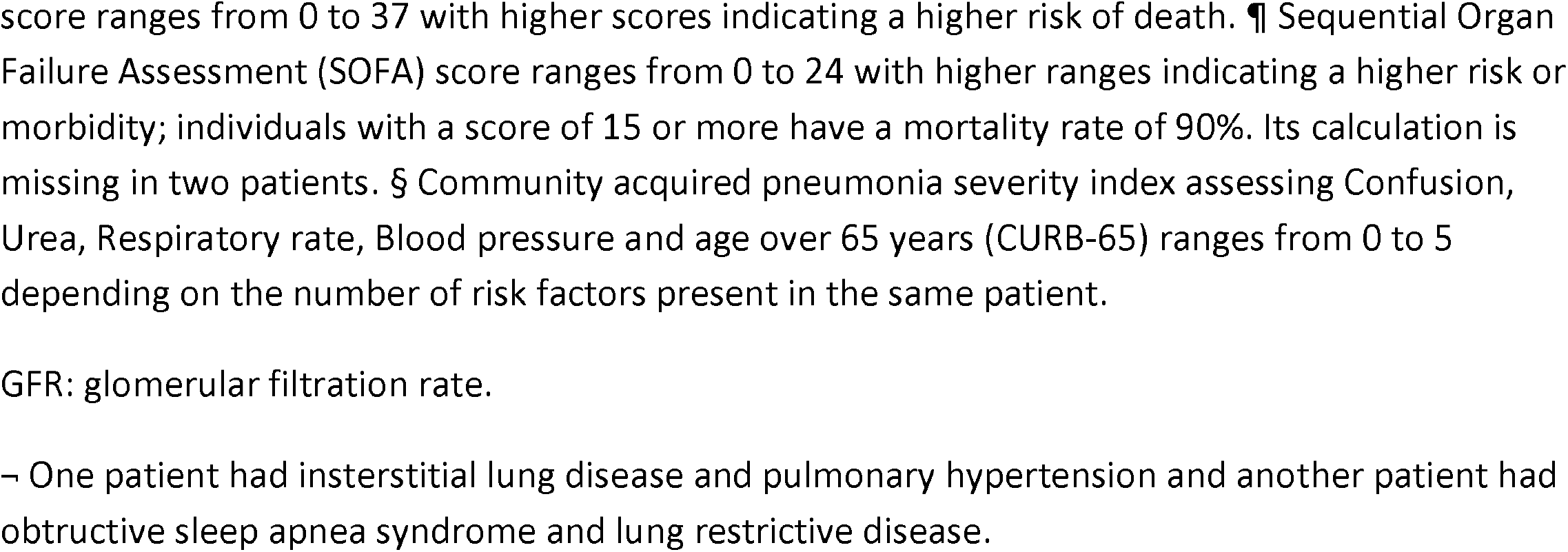
Demographic and clinical characteristics of the patients at baseline*

### Laboratory and Radiologic findings

On admission, mean (±SD) white cell count was 9.2 (10.4) with 17 (21.3%) patients having more than 10000 per cubic millilitre white cells. Lymphocytopenia (<1000 cells per cubic millilitre) was present in 46 (57.5%) patients. Interleukin-6 median (IQR) plasma level on admission was 74.8 (49.4-120.0) ng/ml. Liver enzymes were below five times the upper normal value in all patients. Pneumonia was radiologically proven in all patients on admission or during follow up. Tables 2 and 3 describe laboratory and radiologic findings on admission and during follow up.

**Table 2.**
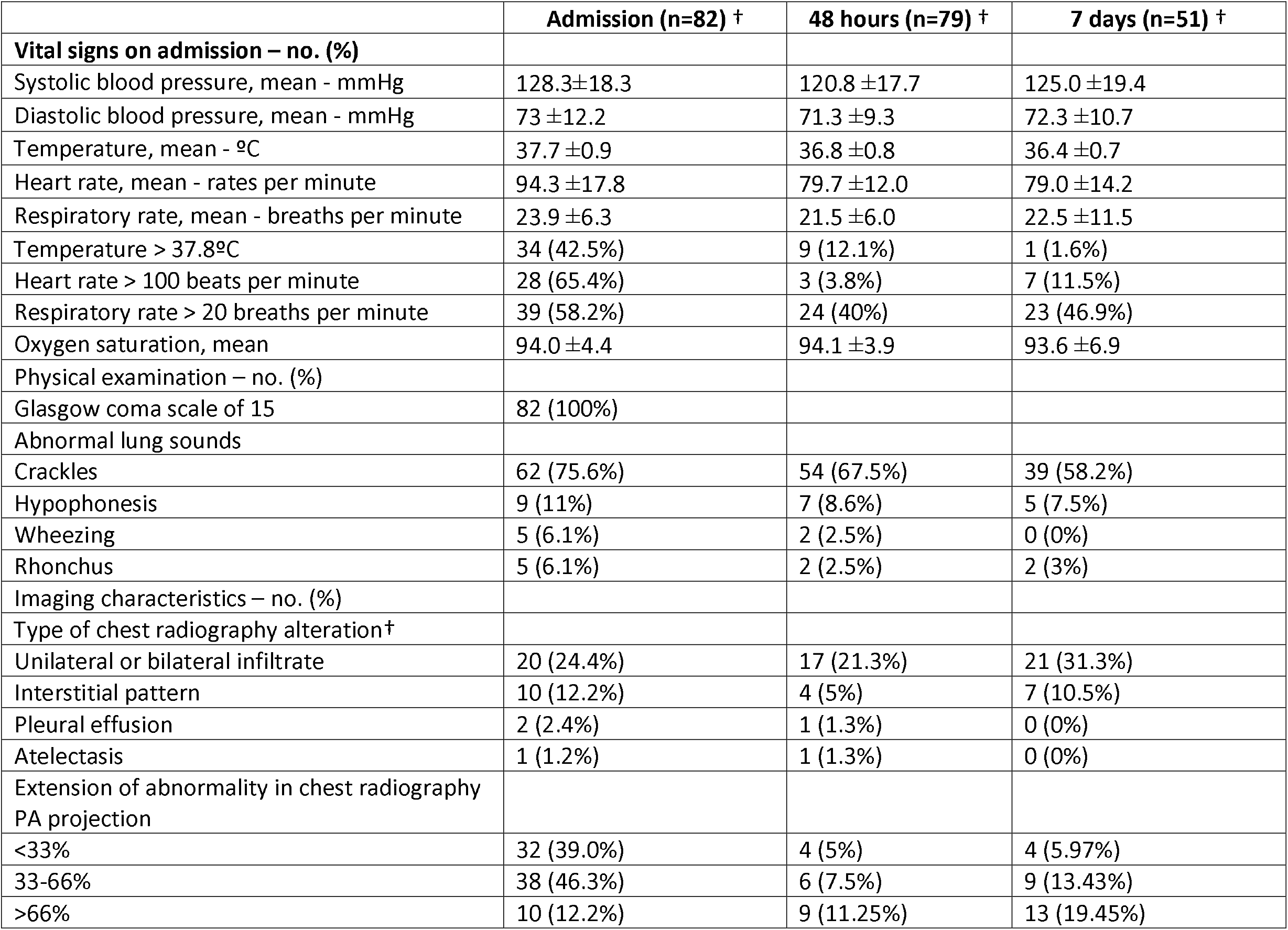
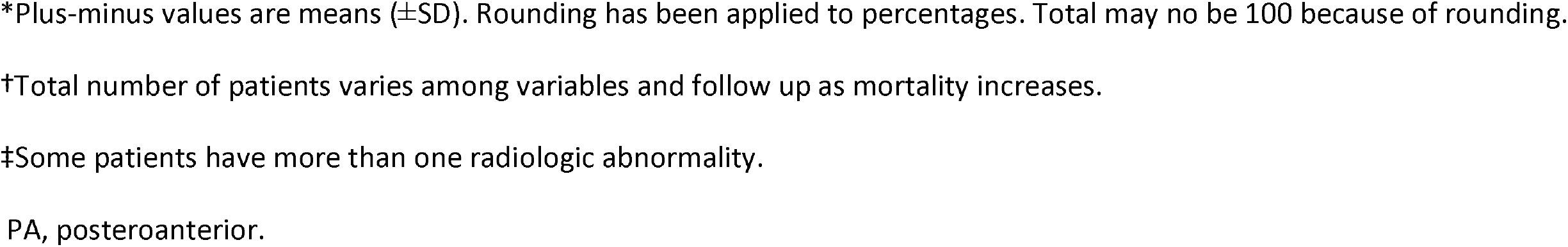
Status on admission and follow up*

**Table 3.** Laboratory data at admission and follow up*

| **Laboratory data – no. (%)** | **Admission (n=82) †** | **48 hours (n=79) †** | **7 days (n=51) †** |
| --- | --- | --- | --- |
| Red cell count |  |  |  |
| Haemoglobin, mean – gr/dl | 13.3 ±1.6 | 12.7 ±1.6 | 12.5 ±1.5 |
| ≥ 10 gr/dl – no. (%) | 78 (96.3%) | 41 (93.2%) | 43 (95.6%) |
| White cell count |  |  |  |
| Mean (SD) – per mm<sup>3</sup> | 9.2 ±10.4 | 6.7 ±3.8 | 7.1 ±3.5 |
| Distribution – no. (%) |  |  |  |
| ≥ 10000/mm<sup>3</sup> | 17 (21.3%) | 5 (11.4%) | 9 (20%) |
| ≤4000/mm<sup>3</sup> | 7 (8.8%) | 10 (22.7%) | 9 (20%) |
| Lymphocyte count |  |  |  |
| Median (IQR) – per mm<sup>3</sup> | 868.9 (593.7-1205.5) | 710.5 (491.5-1154.9) | 1112.0 (575.1-1519.6) |
| Distribution – no. (%) |  |  |  |
| ≥ 1000/mm<sup>3</sup> – no. (%) | 34 (42.5%) | 15 (34.1%) | 23 (51.1%) |
| Platelet count, mean – per mm<sup>3</sup> | 199 ±87.2 | 235.7 ±139.4 | 282.3 ±141.6 |
| Prothrombin time, mean - % | 77.9 ±23.8 |  |  |
| Activated partial thromboplastin time, mean - seconds | 24.6 ±9.8 |  |  |
| Fibrinogen, mean – gr/dl | 5.6 ±1.0 |  |  |
| D dimer, median (IQR) - ng/ml | 295 (201-437) | 565 (303-772) | 738 (273.5-2963) |
| Glucose, mean - mg/dl | 120.2 ±36.7 |  |  |
| Urea, median (IQR) – mg/dl | 43 (38-72) |  |  |
| Serum creatinine, mean - mg/dl | 1.7 ±6.1 | 2.1 ±7.1 | 0.9 ±0.6 |
| Glomerular filtrate, mean – CKD-EPI | 73.5 ±23.8 | 73.1 ±26.5 | 77.25 ±19.9 |
| Sodium, mean – mmol/L | 136.0 ±3.7 |  |  |
| Potassium, mean – mmol/L | 3.9 ±0.7 |  |  |
| Calcium, mean – mg/dl | 8.9 ±0.5 |  |  |
| Total bilirubin, mean – mg/dl | 0.7 ±0.5 |  |  |
| Aspartate aminotransferase, mean - U/litre | 53.1 ±34.3 | 53.7 ±35.4 | 71.4 ±46.2 |
| Aspartate aminotransferase > 40 U/litre | 42 (53.9%) | 25 (61%) | 30 (66.7%) |
| Alanine aminotransferase, mean - U/litre | 41.68 (34.4) | 43.4 (31.7) | 77.3 (71) |
| Alanine aminotransferase > 40 U/litre | 28 (35.4%) | 16 (39.0%) | 30 (66.7%) |
| Alkaline phosphatase, mean - U/litre | 69.80 ±21.7 |  |  |
| Gamma-glutamyl transferase, mean – U/litre | 93 ±58.0 |  |  |
| LDH, mean - UI/L | 446.61 ±79.5 |  |  |
| CRP, mean - mg/dl | 17.98 ±11.7 | 17.5 ±10.0 | 6.3 ±9.2 |
| Ferritin, mean - ng/ml | 885.69 ±500.5 | 1505.4 ±1194.6 | 1241.6 ±789.2 |
| Proteins, mean - gr/dl | 7.38 ±0.7 |  |  |
| Albumin, mean - gr/dl | 3.30 ±0.3 |  |  |
| IL-6, median (IQR) - pg/ml | 74.8 (49.4-120.0) | 184.1 (75.3-592.6) | 501.2 (103.7-2361.0) |
| **Infection analysis** |  |  |  |
| Positive blood cultures | 1 (1.3%) | 0 (0%) | 1 (10%) |
| Positive sputum cultures | 3 (4.3%) | 0 (0%) | 2 (22.2%) |
| Positive pneumococcal urinary antigen | 2 (2.5%) | 0 (0%) | 0 (0%) |

\*Plus-minus values are means (±SD). Rounding has been applied to percentages. Total may no be 100 because of rounding.
†Total number of patients varies among variables and follow up as mortality increases.

### Microbiologic results

All included patients had a positive RT-PCR for SARS-CoV-2 in a respiratory-derived sample. On admission, 2 patients out of 56 had a positive pneumococcal urinary antigen result. Sputum samples from 13 patients were sent on admission, bacterial growth was demonstrated in 3 samples, two with *Haemophilus influenzae* that were considered clinically significant and one was deemed contamination with oral bacteria. During the first 7 days follow up, 2 more positive results were retrieved: one Extended-Spectrum Beta-Lactamase (ESBL)-producing *Escherichia coli* (considered clinically significant) and one *Staphylococcus epidermidis* (considered non-clinically significant). Blood cultures from 65 patients were sent, and one positive bacterial growth (coagulase-negative *Staphylococcus)* was observed, although considered a contamination. Detailed microbiologic data are shown in Table 3.

### Oxygen supplementation and secondary outcomes

Table 4 shows oxygen saturation, oxygen supplementation and ventilation support. On admission, mean FiO2 oxygen supplementation was 0.36 (±0.26) and mean oxygen saturation was 94% (±4.39). Regarding oxygen supplementation devices on admission, 34 (41.5%) patients were on oxygen supplementation: two (5.8%) patients were on nasal cannulas, 22 (64.7%) were using face masks, 9 (26.5%) patients were using high oxygen supplementation devices and 1 (2.9%) patient required endotracheal intubation with mechanical ventilation. Over time, SpO2/FiO2 ratio deteriorated from a median (IQR) of 428 (316.1-454.8) on admission, 271.4 (158.3-361.5) at 48 hours and 230.2 (118.8-346.4) at 7 days follow up (p<0.001). Fifty-five (69.6%) patients required intensive oxygen therapy, including high flow oxygen delivery systems, high flow nasal cannula, non-invasive mechanical ventilation or invasive ventilation during the study period. Median (IQR) days on high flow oxygen delivery systems, high flow nasal cannula, non-invasive mechanical ventilation or invasive ventilation before progression to other intensive oxygen therapy, outcome attainment or data censoring were 2.0 (1.0-3.0), 4.0 (2.0-6.0), 3.0 (2.0-4.0) and 9.0 (9.0-9.0) respectively. The median (IQR) days from admission to first intensive oxygen therapy was 2.0 (1.0-4.5). Only one (1.2%) patient required vasopressor therapy due to hypotension. No patient required renal replacement therapy. Respiratory failure and ARDS developed in 62 (75.6%) and 45 (54.9%) patients, respectively. Median (IQR) days from symptoms to respiratory failure and ARDS were 8 (6.0-11.0) and 8 (5.0-11.0) respectively. Median (IQR) days from admission to respiratory failure and ARDS were 1 (0.0-3.0) and 2 (1.0-4.0) respectively. Secondary outcomes can be found in Table 5.

**Table 4.** Oxygen supplementation and supportive ventilation on admission and follow up*

|  | **Admission (n=82) †** | **48 hours (n=79) †** | **7 days (n=51) †** |
| --- | --- | --- | --- |
| Respiratory frequency, mean – rate per minute | 23.9 ±6.3 | 21.6 ±6 | 22.5 ±11.5 |
| Oxygen saturation, mean | 94 ±4.4 | 94 ±3.9 | 93.6 ±6.9 |
| SpO2/FiO2 ratio, median (IQR) | 428 (316.1-454.8) | 271.4 (158.3-361.5) | 230.2 (118.8-346.4) |
| **Oxygen supplementation and supportive ventilation – no. (%)** |  |  |  |
| Nasal cannula | 2 (5.9%) | 6 (9%) | 4 (7.0%) |
| Face masks | 22 (64.7%) | 33 (49.3%) | 16 (28.1%) |
| High oxygen supplementation device | 9 (26.5%) | 11 (16.4%) | 6 (10.5%) |
| High flow nasal cannulas | 0 (0%) | 10 (14.9%) | 16 (28.1%) |
| Non-invasive mechanical ventilation | 0 (0%) | 2 (3%) | 3 (5.3%) |
| Invasive mechanical ventilation | 1 (2.9%) | 5 (7.5%) | 12 (21.1%) |

\*Plus-minus values are means (±SD). Rounding has been applied to percentages. Total may no be 100 because of rounding.
†Total number of patients varies among variables and follow up as mortality increases.
SpO2/FiO2 ratio: arterial oxygen saturation measured by pulse oximetry to fraction of inspired oxygen

**Table 5.**
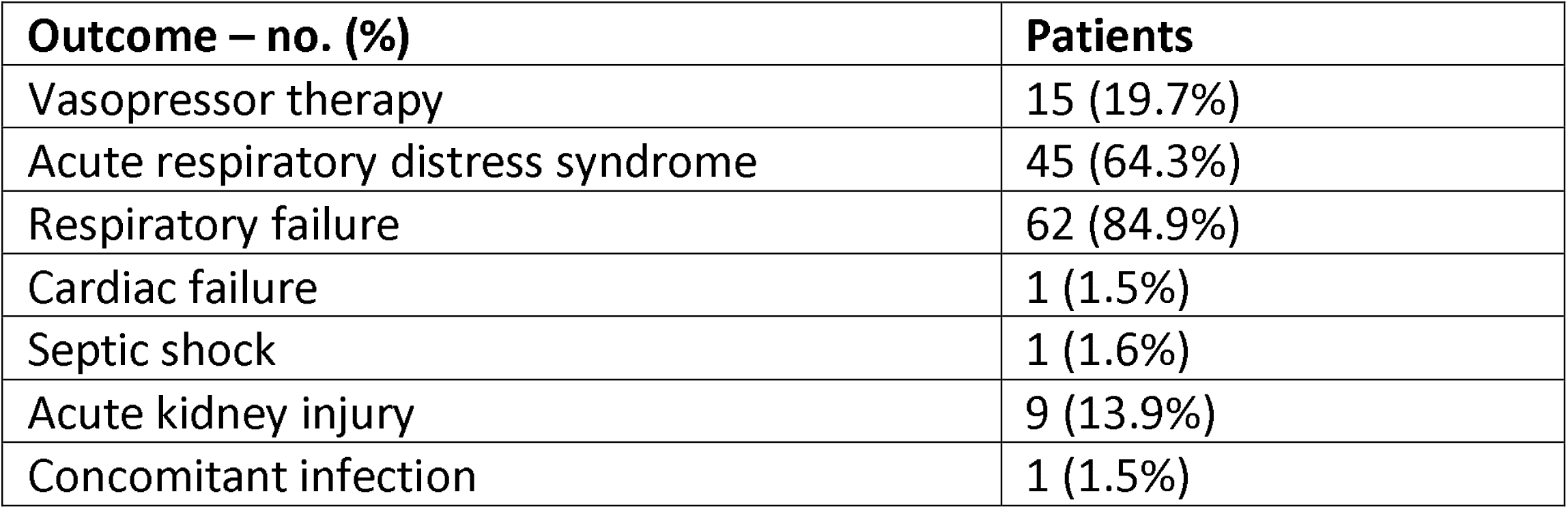
Secondary outcomes at 7-day follow up from tocilizumab administration.

### Tocilizumab treatment and concomitant treatment

Eighty-one (98.9%) patients received hydroxychloroquine, 63 (76.8%) lopinavir/ritonavir, 21 (25.61%) darunavir/cobicistat, and 79 (96.34%) azithromycin. All patients were initially treated with antibiotic therapy, mainly ceftriaxone (77 (93.9%) patients). As expressed before, all patients received at least one dose of tocilizumab. Median (IQR) time in days from symptom onset to tocilizumab administration was 9.0 (6.0-11.0) and from admission to tocilizumab administration was 2.0 (1.0-3.0)). Other treatments include cytokine hemoadsorption therapy in 2 (2.4%) patients in ICU.

### Primary outcome and mortality risk factors

Table 6 summarizes primary outcome in the study population. At the end of the follow up period, of the 82 patients 34 (41.5%) had been discharged, 22 (26.8%) had died, 14 (17.1%) were hospitalized in ICU, 9 (11.0%) were hospitalized in medical wards, and 3 (3.7%) had been transferred to another institution. In the univariate analysis age, age-adjusted Charlson comorbidity index, medical history of active or former solid cancer, hypertension, history of heart failure, chronic kidney disease and worse age-adjusted Charlson index at baseline were associated with increased risk of mortality (Table 7). By 7-day follow-up, the mortality rate was 4.0% per person-day (95% confidence interval [CI], 2.4% to 6.2%) by Kaplan-Meier analysis. Mortality was more frequent in patients receiving tocilizumab once ARDS was present (hazard ratio for mortality 3.3; 95% CI, 1.3 to 8.5; age-adjusted hazard ratio for mortality 2.1; 95% CI, 0.8 to 5.8)(Figure 1) or respiratory failure was present (hazard ratio for mortality 3.13; 95% CI, 1.3 to 7.8; age-adjusted hazard ratio for mortality 2.4; 95% CI, 0.9 to 6.4)(Figure 2). When dividing patients according to the nearest SpO2/FiO2 ratio to tocilizumab administration, mortality was higher among patients with lower SpO2/FiO2 ratio (SpO2/FiO2 ratio<200, 46.2%; SpO2/FiO2 ratio 200-300, 16.7%; SpO2/FiO2 ratio >300, 20.6%; p=0.03)(Figure 3). Distribution of the nearest SpO2/FiO2 ratio to tocilizumab administration depending on outcome groups was not statistically significant (mean (SD) SpO2/FiO2 ratio: 321.3 (154.7) dead, 343.1 (132.7) ICU, 396.9 (96.2) alive; p=0.2)(Figure 4). No correlation was observed between nearest IL-6 levels to tocilizumab administration and main outcome (median (IQR) IL-6 levels: 79.7 (48.2-128.1) dead, 77.5 (55-120) ICU admission, 71.4 (49.4-116) alive; p=0.92). Basal characteristics of patients stratified by ARDS and respiratory failure can be found in the Supplementary Appendix.

**Table 6.** Main outcome at 7-day follow up from tocilizumab administration.

| **Main outcomes – no. %** | **Patients** |
| --- | --- |
| Discharge | 34 (41.5%) |
| In-patient in conventional ward | 9 (11.0%) |
| Intensive care unit | 14 (17.1%) |
| Death | 22 (26.8%) |
| Transferred | 3 (3.7%) |

**Table 7.** Comparison of risk factors by in-hospital mortality.

| Characteristics | Alive (n=60) | Dead (n=22) | P-value |
| --- | --- | --- | --- |
| Demographics |  |  |  |
| Age, mean (SD) - yr | 53.3 (19.9) | 75.2 (6.2) | <0.001 |
| Sex – no (%) | 36 (60.0) | 16 (72.7) | 0.289 |
| Coexisting condition – no (%) |  |  |  |
| Tobacco use |  |  | 0.397 |
| Active tobacco smoker | 5 (8.3) | - |  |
| Former tobacco smoker | 19 (31.7) | 9 (40.9) |  |
| Never smoke | 36 (60.0) | 13 (59.1) |  |
| Alcohol use |  |  | 0.070 |
| Active daily alcohol consumption | 1 (1.7) | - |  |
| Former daily alcohol consumption | - | 2 (9.1) |  |
| Never drink daily | 59 (98.3) | 20 (90.9) |  |
| Barthel index at admission, median (IQR) | 100 (100-100) | 100 (100-100) | 0.371 |
| Dementia | 1 (1.7) | - | 1 |
| Diabetes Mellitus | 12 (20.0) | 4 (18.2) | 1 |
| Immunosuppression | 5 (8.3) | 5 (22.7) | 0.123 |
| Solid tumor | 3 (5.1) | 6 (27.3) | 0.012 |
| Leukemia/Lymphoma | 2 (3.3) | 1 (4.6) | 1 |
| Hypertension | 17 (28.3) | 15 (68.2) | 0.001 |
| Chronic heart failure | 1 (1.7) | 4 (18.2) | 0.017 |
| Chronic lung disease | 11 (18.6) | 8 (36.4) | 0.094 |
| Chronic renal failure | 4 (6.7) | 7 (33.3) | 0.005 |
| Liver cirrhosis | 1 (1.7) | - | 1 |
| Central nervous system disease | 1 (1.7) | 1 (4.6) | 0.467 |
| Obesity | 14 (23.3) | 3 (13.6) | 0.539 |
| Median (IQR), age-adjusted Charlson index at baseline - points | 2 (1 – 3) | 5 (3 – 6) | <0.001 |
| Oxygenation previous to tocilizumab administration |  |  |  |
| Oxygen saturation (pulse oximeter) at baseline, median (IQR) | 95 (94 - 97.5) | 93 (89 - 97) | 0.061 |
| FiO2 at baseline, median (IQR) | 0.21 (0.21 - 0.28) | 0.26 (0.21 - 0.50) | 0.130 |
| SpO2/FiO2 ratio at baseline, median (IQR) | 440 (343-455) | 393 (180-452) | 0.134 |
| High oxygen supplementation or ventilation at baseline* - no. (%) | - | 1 (4.6%) | 0.268 |
| Oxygen saturation (pulse oximeter) at tocilizumab administration, median (IQR) | 95 (93-97) | 92 (89-94) | 0.006 |
| FiO2 previous to tocilizumab administration, median (IQR) | 0.27 (0.21-0.40) | 0.35 (0.21-1) | 0.131 |
| SpO2/FiO2 ratio previous to tocilizumab administration, median (IQR) | 354 (228-438) | 263 (95-423.8) | 0.072 |
| High oxygen supplementation or ventilation previous to tocilizumab administration* | 3 (5%) | 4 (18%) | 0.79 |
| Days from initial symptoms to tocilizumab administration, median (IQR) | 9 (7 - 11) | 7 (5 - 15) | 0.372 |
| Days from admission to tocilizumab administration, median (IQR) | 2 (1 - 3) | 3 (1 - 4) | 0.064 |
| Days from respiratory insufficiency to tocilizumab administration, median (IQR) | 0 (0 - 1) | 1 (0 - 2) | 0.055 |
| Days from ARDS to tocilizumab administration, median (IQR) | 0 (-1 - 0) | 0 (0 - 1) | 0.132 |

\*Plus-minus values are means ( ± SD). Rounding has been applied to percentages. Total may not be 100 because of rounding.
†Includes high flow oxygen delivery systems, high flow nasal cannula, non-invasive mechanical ventilation or invasive ventilation
ARDS: acute respiratory distress syndrome.

### Safety

Twelve (14.63%) out of 82 patients reported a total of 14 adverse events during the follow up. Thirteen (92.9%) adverse events were considered related to lopinavir/ritonavir, 9 (75.0%) patients discontinued lopinavir/ritonavir treatment due to gastrointestinal symptoms. Diarrhoea was the most common reported adverse event. Other adverse events included nausea and dysuria. There were no adverse events attributed to tocilizumab. No serious adverse events were reported during follow up, and only 2 (14.3%) were considered moderate. Eleven (91.7%) patients recovered without medical sequelae and one patient had an unknown outcome. No tocilizumab discontinuation was reported due to adverse events.

## DISCUSSION

This preliminary report from the Vall d’Hebron COVID-19 Prospective Cohort Study describes the characteristics and clinical outcomes of patients who were hospitalized in non-ICU wards and received treatment with tocilizumab. Our results show that a timely administration of immune-modulating therapies, before the onset of respiratory insufficiency or ARDS, may improve severe COVID-19 patients’ outcomes. Therapies to improve outcomes of patients with COVID-19 focus on viral-directed therapies and host-directed therapies. There is still lack of evidence about the efficacy of any of these therapies, although this does not prevent physicians to use all sorts of off-label therapies despite the risk of serious adverse events.^26^ Therapies to curb uncontrolled cytokine release have been proposed and are being widely used. Randomized controlled trials with host-directed therapies are currently under way and results will be available within the next months.^17,18^ Meanwhile, data from prospective studies can help to improve COVID-19 patient management. In our study, the 7-day mortality was 26.8%, slightly higher than a recent experience with remdesivir and similar to the mortality of 22.1% reported in another study with lopinavir/ritonavir.^27,28^ However, the patients in our prospective cohort had more coexisting conditions, including potential mortality risk factors such as hypertension, other cardiovascular and metabolic diseases, chronic kidney disease and cancer.

The understanding of mortality risk factors in patients with COVID-19 is an evolving matter. Age, specific coexisting conditions and laboratory parameters may help identify patients with poor outcome.^29^ As expressed before, in our cohort of patients treated with tocilizumab, hypertension, history of cardiac failure and chronic kidney disease were associated with higher mortality in the univariate analysis. Antihypertensive agents, such as angiotensin-converting enzyme inhibitors (ACEI) and angiotensin receptor blockers (ARB), have been suggested to be associated with the increased mortality observed in this subset of patients. Angiotensin converting enzyme 2 plays an important role in SARS-CoV-2 viral entry as co-receptor.^30^ Hypothesis outline that the interactions between these drugs and co-receptors may increase viral spreading in the lung and increase risk of death. However, the evidence is limited and no specific recommendations could be drawn from current evidence, especially when ACEI and ARB have shown to reduce mortality in this at risk population.^31^ The Vall d’Hebron COVID-19 Prospective Cohort Study has among its main objectives to analyse the role of these and other drugs used to treat chronic conditions in the prognosis of patients with COVID-19.

Host-directed therapies aiming at blocking an unrestrained immune response and an excessive inflammation have been proposed as potential therapies to prevent acute lung injury and subsequent ARDS. SARS-CoV-2 infection severity has been associated with an increase in IL-6 and D dimer levels, and the cytokine profile resembles that of other conditions in which host-directed therapies have been successfully used.^13,29,32^ Timely use of host-directed therapies may curb uncontrolled cytokine release and prevent damage inflicted by hyperinflammation. Tocilizumab has shown to be safe in two small Chinese cohort studies of patients with COVID-19.^9,16^ Efficacy of tocilizumab, measured as hospital discharge, was 90.5% (19 out of 21 patients) in patients with COVID-19 needing oxygen supplementation and having elevated IL-6 in one study^9^; while the other study showed 3 deaths, 10 clinical stabilizations and 2 clinical aggravations out of 15 patients.^16^ Other laboratory and radiologic findings also improved in a short period of time. However, standard of care in the studies was different limiting the possibility of direct comparisons. As in other infections and inflammation-driven diseases, timely initiation of precise therapy is the mainstay of patient management and directly affects mortality and morbidity. Our study showed that patients receiving prompt treatment with tocilizumab before lung injury is established have less 7-day mortality, a benefit that may be sustained in the long-term.^33^ The safety profile of tocilizumab has been extensively studied in clinical trials with patients suffering autoimmune diseases. The most common adverse events of intravenous tocilizumab in a pooled analysis of 3 clinical trials were upper respiratory infections, nasopharyngitis, headache, hypertension and increase in liver enzymes. Serious adverse events occurred in 12% of the patients, infectious diseases being the most common.^34^ In our study we did not report any serious adverse events, although the symptoms of systemic viral infections may mimic any adverse event and make its identification difficult. Tozilizumab-related bacterial infections were not reported in our study. Two factors may have contributed to this: first, many patients were under antibiotic treatment, and second, the short follow up period precludes us from any further analysis. Nevertheless, the low cumulative dose administered in this subset of patients may diminish the likelihood of infectious adverse events.

In a time of scarce medical resources, including limited stock of host-directed therapies, hard medical decisions have to be done by front-line physicians. Allocation of therapies to patients with the highest chances of a favourable outcome should be encouraged, maximizing the benefit of the intervention.^35^ Evidence-based decision-making and benefit-maximizing allocation of the available resources should be promoted. In this regard our study can help physicians to better allocate host-directed therapy in patients with COVID-19 prioritizing moderate-to-severe ill patients over critically ill patients.

This preliminary exploratory study has several limitations. First, there is no control group and the minimum follow up period was 7 days. Therefore, at this point it is not possible to evaluate the differences between patients receiving tocilizumab or not and, consequently, it is not possible to evaluate solidly what is the overall benefit of administering this drug. However, the urgency of obtaining data on new therapies justifies the early communication of these results. Second, ICU admission is not a very robust endpoint since it depends on the attitudes of the treating physicians as well as the availability of beds at times of resource scarcity and overwhelming demand. For this reason, mortality was selected as the main outcome in our study. Besides, the subsequent analysis of all patients included in the Vall d’Hebron COVID-19 Prospective Cohort Study may solve this limitation and inform results with a longer follow up period. Finally, our data is limited to a single centre. While our results may not be extrapolated to other populations or other standards of care, the management of patients was homogeneous avoiding the centre effect of multicentric studies. Multivariate analysis is limited by sample size.

## CONCLUSION

In summary, we found a mortality rate of 26.8% in this subset of patients with COVID-19 receiving tocilizumab for the treatment of inflammatory-related lung injury. Time from lung injury onset to tocilizumab administration may be critical to patient recovery. Our results may help front-line physician to make evidence-based decisions in times of scarce resources and operationalized fair and transparent allocation procedures, maximizing the benefit of the intervention. Future and current host-directed clinical trials for patients with COVID-19 should consider our preliminary data in their design. All our patients were treated with a combination of antiviral drugs whose efficacy is yet to be demonstrated. Host-directed therapies in the absence of antiviral drugs needs further investigation.

## Data Availability

Unidentified dataset can be made available upon request to the corresponding author (ASM) after publication of the manuscript.

## ACKNOWLEGDEMENT

None

## CONTRIBUTION STATEMENT

- Conceived the idea: ASM, JEP, JSN, NFH, BA
- Reviewed medical chart and collected data. ASM, JSM, JEP, NFH, FS, XD, MM, AA, SEE, ASG, AMP, PBM, SA, JS, AGdC.
- Supervised the findings: ASM, JEP, JSN, NFH, BA
- Performed statistical analysis: ASM, JEP, NFH, SPH
- Wrote the manuscript with inputs from all authors: ASM, JEP, NFH
- Supervised the project: ASM, JS, BA
- Interpretation of the results: ASM, JSM, JEP, NFH, FS, XD, MM, AA, SEE, ASG, AMP, PBM, SA, JS, AGdC.
- Critically reviewed the manuscript: ASM, JSM, JEP, NFH, FS, XD, MM, AA, SEE, ASG,
- AMP, PBM, SA, JS, AGdC.

## FIGURES

Figure 1. 7-day mortality curves according to the moment patients received tocilizumab: before or after developing ARDS. ARDS: acute respiratory distress syndrome.

Figure 2. 7-day mortality curves according to the moment patients received tocilizumab: before or after developing respiratory insufficiency.

Figure 3. Outcome according to nearest SpO2/FiO2 ratio to tocilizumab administration Figure 4. Nearest SpO2/FiO2 ratio to tocilizumab administration distribution according to 7-days outcome

## REFERENCES

1. Huang C, Wang Y, Li X, et al. Clinical features of patients infected with 2019 novel coronavirus in Wuhan, China. Lancet Lond Engl 2020;395(10223):497–506.

2. Novel Coronavirus (2019-nCoV) situation reports [Internet]. [cited 2020 Apr 1];Available from: https://www.who.int/emergencies/diseases/novel-coronavirus-2019/situation-reports

3. Novel Coronavirus Pneumonia Emergency Response Epidemiology Team. [The epidemiological characteristics of an outbreak of 2019 novel coronavirus diseases (COVID-19) in China]. Zhonghua Liu Xing Bing Xue Za Zhi Zhonghua Liuxingbingxue Zazhi 2020;41(2):145–51.

4. Hauser A, Counotte MJ, Margossian CC, et al. Estimation of SARS-CoV-2 mortality during the early stages of an epidemic: a modelling study in Hubei, China and northern Italy. medRxiv 2020;2020.03.04.20031104.

5. De Clercq E. Potential antivirals and antiviral strategies against SARS coronavirus infections. Expert Rev Anti Infect Ther 2006;4(2):291–302.

6. Al-Tawfiq JA, Memish ZA. Update on therapeutic options for Middle East Respiratory Syndrome Coronavirus (MERS-CoV). Expert Rev Anti Infect Ther 2017;15(3):269–75.

7. Gautret P, Lagier J-C, Parola P, et al. Hydroxychloroquine and azithromycin as a treatment of COVID-19: results of an open-label non-randomized clinical trial. Int J Antimicrob Agents 2020;105949.

8. Chen Z, Hu J, Zhang Z, et al. Efficacy of hydroxychloroquine in patients with COVID-19: results of a randomized clinical trial. medRxiv 2020;2020.03.22.20040758.

9. Xu X, Han M, Li T, et al. Effective treatment of severe COVID-19 patients with tocilizumab. Available from: chinaXiv:202003.00026v1 (2020). www.chinaxiv.org/abs/202003.00026

10. Tang N, Bai H, Chen X, Gong J, Li D, Sun Z. Anticoagulant treatment is associated with decreased mortality in severe coronavirus disease 2019 patients with coagulopathy. J Thromb Haemost JTH 2020;

11. Richardson P, Griffin I, Tucker C, et al. Baricitinib as potential treatment for 2019-nCoV acute respiratory disease. Lancet Lond Engl 2020;395(10223):e30–1.

12. Mehta P, McAuley DF, Brown M, Sanchez E, Tattersall RS, Manson JJ. COVID-19: consider cytokine storm syndromes and immunosuppression. The Lancet 2020;395(10229):1033–4.

13. Zumla A, Hui DS, Azhar EI, Memish ZA, Maeurer M. Reducing mortality from 2019-nCoV: host-directed therapies should be an option. Lancet Lond Engl 2020;395(10224):e35–6.

14. Biggioggero M, Crotti C, Becciolini A, Favalli EG. Tocilizumab in the treatment of rheumatoid arthritis: an evidence-based review and patient selection. Drug Des Devel Ther 2019;13:57–70.

15. Brudno JN, Kochenderfer JN. Toxicities of chimeric antigen receptor T cells: recognition and management. Blood 2016;127(26):3321–30.

16. Luo P, Liu Y, Qiu L, Liu X, Liu D, Li J. Tocilizumab treatment in COVID-19: a single center experience. J Med Virol 2020;

17. Evaluation of the Efficacy and Safety of Sarilumab in Hospitalized Patients With COVID-19 - Full Text View - ClinicalTrials.gov [Internet]. [cited 2020 Apr 11];Available from: https://clinicaltrials.gov/ct2/show/NCT04315298

18. Study of the Efficacy and Safety of Ruxolitinib to Treat COVID-19 Pneumonia - Full Text View - ClinicalTrials.gov [Internet]. [cited 2020 Apr 11];Available from: https://clinicaltrials.gov/ct2/show/NCT04331665

19. Lim WS, van der Eerden MM, Laing R, et al. Defining community acquired pneumonia severity on presentation to hospital: an international derivation and validation study. Thorax 2003;58(5):377–82.

20. Seymour CW, Liu VX, Iwashyna TJ, et al. Assessment of Clinical Criteria for Sepsis. JAMA 2016;315(8):762–74.

21. World Health Organization. Laboratory testing for 2019 novel coronavirus (2019-nCoV) in suspected human cases. WHO - Interim Guid 2020;2019(January):1–7.

22. Moës E, Vijgen L, Keyaerts E, et al. A novel pancoronavirus RT-PCR assay: frequent detection of human coronavirus NL63 in children hospitalized with respiratory tract infections in Belgium. BMC Infect Dis 2005;5:6.

23. Woo PCY, Lau SKP, Lam CSF, et al. Discovery of a novel bottlenose dolphin coronavirus reveals a distinct species of marine mammal coronavirus in Gammacoronavirus. J Virol 2014;88(2):1318–31.

24. Force TADT. Acute Respiratory Distress Syndrome: The Berlin Definition. JAMA 2012;307(23):2526–33.

25. Festic E, Bansal V, Kor DJ, Gajic O. SpO2/FiO2 Ratio on Hospital Admission Is an Indicator of Early Acute Respiratory Distress Syndrome Development Among Patients at Risk. J Intensive Care Med 2013;30(4):209–16.

26. Kalil AC. Treating COVID-19-Off-Label Drug Use, Compassionate Use, and Randomized Clinical Trials During Pandemics. JAMA 2020;

27. Grein J, Ohmagari N, Shin D, et al. Compassionate Use of Remdesivir for Patients with Severe Covid-19. N Engl J Med 2020;

28. Cao B, Wang Y, Wen D, et al. A Trial of Lopinavir-Ritonavir in Adults Hospitalized with Severe Covid-19. N Engl J Med 2020;

29. Zhou F, Yu T, Du R, et al. Clinical course and risk factors for mortality of adult inpatients with COVID-19 in Wuhan, China: a retrospective cohort study. Lancet Lond Engl 2020;395(10229):1054–62.

30. Zhou P, Yang X-L, Wang X-G, et al. A pneumonia outbreak associated with a new coronavirus of probable bat origin. Nature 2020;579(7798):270–3.

31. Patel AB, Verma A. COVID-19 and Angiotensin-Converting Enzyme Inhibitors and Angiotensin Receptor Blockers: What Is the Evidence? JAMA 2020;

32. Lee DW, Gardner R, Porter DL, et al. Current concepts in the diagnosis and management of cytokine release syndrome. Blood 2014;124(2):188–95.

33. Ngai JC, Ko FW, Ng SS, To K-W, Tong M, Hui DS. The long-term impact of severe acute respiratory syndrome on pulmonary function, exercise capacity and health status. Respirol Carlton Vic 2010;15(3):543–50.

34. Nishimoto N, Miyasaka N, Yamamoto K, Kawai S, Takeuchi T, Azuma J. Long-term safety and efficacy of tocilizumab, an anti-IL-6 receptor monoclonal antibody, in monotherapy, in patients with rheumatoid arthritis (the STREAM study): evidence of safety and efficacy in a 5-year extension study. Ann Rheum Dis 2009;68(10):1580–4.

35. Emanuel EJ, Persad G, Upshur R, et al. Fair Allocation of Scarce Medical Resources in the Time of Covid-19. N Engl J Med 2020;

